# Planning phase-2 for the endoscopic units in Northern Italy after COVID-19 lockdown: an exit strategy with a lot of critical issues and a few opportunities

**DOI:** 10.1101/2020.05.12.20092270

**Authors:** Gianpiero Manes, Alessandro Repici, Franco Radaelli, Simone Saibeni, Cristina Bezzio, Matteo Colombo, Fabio Pace, on behalf of The ITALIAN GI-COVID19 Working Group, Mario Schettino, Massimo Devani, Paolo Andreozzi, Lucienne Pellegrini, Desirè Picascia, Arnaldo Amato, Costanza Alvisi, Giovanni Aragona, Elia Armellini, Mohammad Ayoubi, Stefano Bargiggia, Stefano Benvenuti, Paolo Beretta, Paola Boarino, Andrea Buda, Lorenzo Camellini, Sergi Cavenati, Vincenzo Cennamo, Fabrizio Cereatti, Claudio de Angelis, Giovanni de Pretis, Giuseppe De Roberto, Marco Dinelli, Giulio Donato, Carlo Fabbri, Luca Ferraris, Alessandro Fugazza, Pietro Fusaroli, Nicola Gaffuri, Salvatore Greco, Piera Leoni, Gianluigi Longobardi, Benedetto Mangiavillano, Mauro Manno, Alberto Merighi, Guido Missale, Alessandro Mussetto, Massimiliano Mutignani, Pietro Occhipinti, Roberto Penagini, Raffaele Salerno, Romano Sassatelli, Sergio Segato, Sandro Sferrazza, Teresa Staiano, Alberto Tringali, Giovanna Venezia, Carlo Verna

## Abstract

**Background and aims:** Restarting activity in Endoscopic Departments (ED) after COVID-19 lockdown raises critical issues. This survey investigates strategies and uncertainties on resumption of elective activity.

**Methods:** Directors of 55 EDs in Northern Italy received a questionnaire focusing on the impact of pandemic on activity and organization and on the resources available at re-opening. A section was devoted to gather forecasts and proposals on the return path to normality.

**Results:** All centres had reduced their activities of at least 50% of the pre-COVID-19 period. A rate of endoscopists (13.6%), nurses (25.2%), and health assistants (14%) were not available since infected, or relocated to other departments. One third of endoscopic rooms were converted to COVID-19 care. Two third had the waiting or the recovery areas too small for distancing. A dedicated pathway for infected patients could not be guaranteed in 20% of EDs. Only one third of EDs judged realistic to completely restore a pre-crisis workload by the next months. Optimizing appropriateness of procedures, closer interaction with GPs and triaging patients with telemedicine were the proposals to re-open EDs.

**Conclusions:** The critical issues while re-opening EDs calls for reducing the workload in the endoscopy units through appropriate rescheduling of procedures.

**Funding:** None

## Background

Since the beginning of May, Italy will gradually get out of the COVID-19 lockdown^1^. The third-largest economy in the Eurozone had been under lockdown measures since March 9^th^, in order to contain the viral outbreak. With the deceleration of the epidemic, a reduction of of constraint measures, allowing certain businesses activities to progressively reopen has taken place through a well-structured and articulated plan and Italy is now in the so called “phase-2” of epidemic, an intermediary period between the current strict lockdown and “phase-3”, when the country should gradually return to normality. After a pause for all but emergency and essential procedures in the acceleration phase of current epidemic, Endoscopy Departments (ED) are now reconverting to their pre-COVID-19 configuration and progressively returning to elective endoscopy^2–4^.

However, restarting safely the out patient clinic activity raises several critical issues, such as the high risk of healthcare personnel and patient exposure to the COVID-19 infection, the reduced availability of healthcare personnel, the presence of infrastructural barriers (i.e. inadequate spaces to guarantee the safe distancing in both waiting and recovery rooms), the lack of a clear and thoughtful policy regarding the timely rescheduling of cancelled or postponed endoscopies during the lockdown.

The aim of present survey is to investigate among the EDs located in Northern Italy – the mostly hit area by the COVID-19 outbreak all over Europe - potential barriers and strategies to safely resume the elective endoscopy activity on the way out of the current lockdown.

## Methods

Due to the study design, our ethical committee waived ethical approval. The study was conducted as a survey between April 20^th^ and April 25^th^, 2020 (about two weeks before the start of lockdown “phase-2”). Directors of EDs in high-risk areas of Northern Italy were invited by e-mail to complete a structured questionnaire.

A total of 55 centres, all participating in a previous survey on the burden of COVID-19 outbreak of EDs in Italy (the ITALIAN GI-COVID19 Working Group) (3), were invited. No incentive was offered for participation.

The questionnaire is reported as Supplementary Table. It was not anonymous and consisted of 3 sections with mostly multiple-choice questions. Briefly, the first section focused on organizational characteristics of EDs before and after the COVID-19 outbreak, in order to assess its impact of epidemic on endoscopy services. Some infrastructural characteristics of EDs, which were considered to be crucial for limiting the infection exposure of healthcare personnel, patients and accompanying visitors, were also collected. The second section explored the availability of specialist staff (endoscopists, nurses and assistants) and personal protective equipment (PPE) in the EDs at the start of phase-2. In the third section, the directors of each ED were asked to foresee the endoscopy workload they would realistically estimate as doable, according to the local resources, in the early recovery phase of COVID-19 pandemic (since May to July 2020), and to indicate strategies to optimize endoscopic activity.

### Statistics

Categorical data are presented as proportion, whereas continuous data as mean and standard deviation (SD) or, in case of skewed distribution, as median and interquartile range (IQR).

## Results

Of 55 invited EDs, 43 (78.2%) participated and completed the questionnaire. Of them, 40 were high-volume centres, performing > 5000 procedures/yr. The median interval time (IQR) from construction or last renovation of EDs was 9·8 (7·2-13·2) (range 1-25 years). The main characteristics of the EDs in the pre-COVID-19 period and at the time of questionnaire administration, just before the beginning of phase-2, are shown in Table 1. Due to the COVID-19 outbreak, 17 (39·5%) centres had their normal endoscopic activities reduced by 50-74% and the remaining 26 (60·5%) by 75-99%.

**Table 1:** The main characteristics of the Endoscopic Departments in the pre-COVID-19 period and at the time of questionnaire administration ED: Endoscopic Department

|  |  |
| --- | --- |
| **Involved Endoscopic Departments (n)** | 43 |
| **EDs performing >5000 exams/year (n, rate)** | 40/43 (93%) |
| **Time from ED construction or renovation (year; mean ± SD; range)** | 11.9 ± 7.3; 1-25 |
| **EDs >10 year-old (n, rate)** | 20/43 (46.5%) |
| **Available/non-available endoscopic rooms** | 121/67 |
| **EDs with at least one non-available endoscopic room (n, rate)** | 32/43 (74.4%) |
| **Available endoscopic rooms/ED (mean ± SD; range)** |  |
| - Pre-COVID-19 | 4.44 ± 2.13; 2-11 |
| - Present time | 2.80 ± 1.38; 1-7 |
| **COVID-19 -related procedure reduction (n, rate)** |  |
| - 50-74% | 17/43 (39.5%) |
| - 75-99% | 26/43 (60.5%) |
| **Available endoscopists/ED (mean ± SD; range)** |  |
| - Pre-COVID-19 | 8.34 ± 4.53; 3-20 |
| - Present time | 7.27 ± 4.51; 1-19 |
| **Available/non-available endoscopists (n)** | 305/48 |
| **Reason for non-availability** |  |
| - COVID-19 infection | 9 |
| - Reallocation to another unit | 39 |
| **Available nurses/ED (mean ± SD; range)** |  |
| - Pre-COVID-19 | 15.19 ± 10.46; 3-65 |
| - Present time | 11.34 ± 9.92; 2-62 |
| **Available/non-available nurses** | 481/162 |
| **Reason for non-availability** |  |
|  | 25 |
| - COVID-19 infection<br>- Reallocation to another unit | 137 |
| **Available health assistant/ED** (mean ± SD; range)<br>- Pre-COVID-19<br>- Present time | 4.24 ± 3.36; 1-15<br>3.22 ± 2.17; 0-15 |
| **Available/non-available health assistants**<br>Reason for non-availability<br>- COVID-19 infection<br>- Reallocation to another unit | 154/25<br>4<br>21 |

### Staff requirements of the endoscopic departments

Overall, 353 endoscopists (range 3-20/ED), 643 nurses (range 7-65/ED), and 179 health assistants (range 1-15/ED) had been working in the services in the pre-COVID-19 period. At the end of April, 48/353 (13·6%) endoscopists, 162/643 (25·2%) nurses, and 25/179 (14%) health assistants were not available since infected (9, 25 and 4 respectively), or reallocated to other COVID units (39, 137 and 21 respectively) (Table 1).

### Structural requirements of the endoscopic departments

Of 188 endoscopy rooms used in the pre-COVID period, 67 (35%) in 32 EDs were not available, since either converted to COVID-19 care area (n=9) or exclusively devoted to endoscopic procedures in COVID-19 patients (n=58). As far as the EDs structural characteristics are concerned, 29 (67·4%) centres had either the waiting area (22/43, 51·2%), or the recovery area (24/43, 55·8%) or both (17/43, 39·5%) too small to guarantee, at the pre-crisis workload, the minimal distance between patients or caregivers/escorts; 10 (23·2%) centres could not guarantee a “patients infected pathway” separated by “noninfected” areas in order to avoid crossing of COVID-19 positive and negative patients and minimize contamination; 30 (70%) were lacking at least one negative-pressure room; 10 (23·2%) did not guarantee even the separated dirty/clean pathways for the endoscopes. In this phase of epidemic, PPE shortage represented a critical issue for a minority of centres, as only 5 (11·6%) centres reported to be still suffering from it. In general, only 3 (7%) EDs reported to be able to resume the elective endoscopic activity at the pre-crisis volume with respect of all the safety prescriptions.

### Future perspectives and proposals

We asked the directors of participating centres to foresee which increase in the endoscopic workload would have been bearable in the upcoming months of the recovery phase, according to the services resources (Figure 1). For the month of May, the majority of them (34/43, 79%) envisioned as realistic a workload increase up to 33%, as compared to the actual one. For the month of June, this figure was up to 33% and 50% in 26 centres (60·5%) and in 15 (34·9%), respectively. For the month of July, the majority of centres (30/43, 69·8%) envisioned a workload increase of at least 50% (n=22). Returning to the pre-crisis workload by the end of July, September, and October was judged as a realistic goal by 8 (18·6%), 10 (23·2%), and 14 (32·6%) participants, respectively.

**Figure 1:**
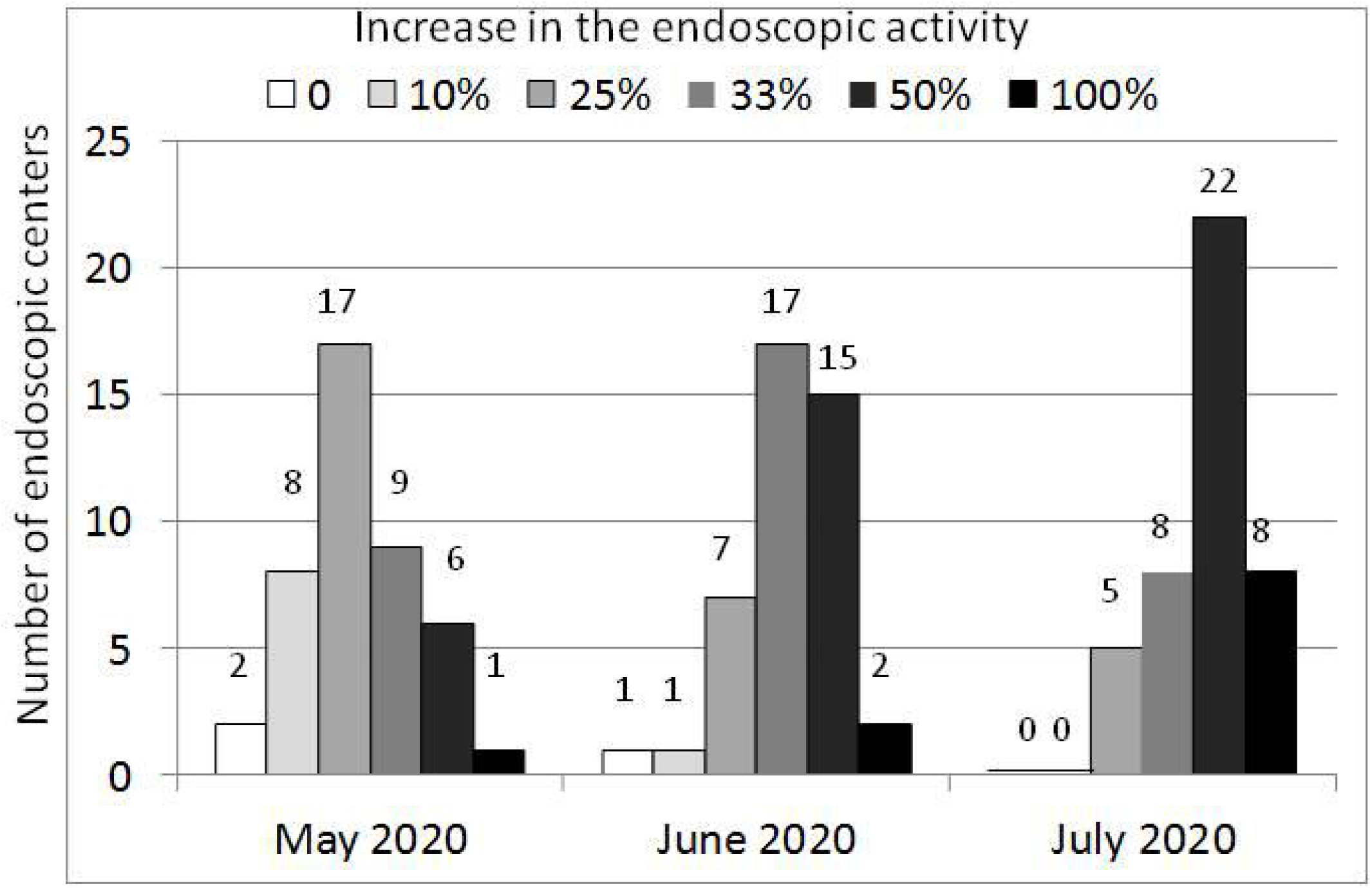
Hypothesized increase in the endoscopic workload, as compared to the actual one, bearable in the upcoming months after re-opening of the Endoscopic Departments

All participants agreed that, once the EDs will have completely returned to elective endoscopy, the chance of overcrowding would be very high, due to the huge number of postponed cases to be rescheduled. In spite of that, only 10 of them reported to be confident in being able to significantly increase (at least 33%) their activity with respect to the pre-COVID-19 volume; conversely, the majority of participants realistically declared no (14 centres) or minimal (10-25%) increases in their activities to be possible.

All endoscopists hypothesized that future strategies to optimize endoscopic activity would have necessarily implied a more rational use of the endoscopic resources minimizing inappropriate procedures; they suggested that this could be achieved by implementing the application of current guidelines (n=12), organizing a direct-line among general practitioners (GP) and endoscopists to facilitate triaging and scheduling/rescheduling patients (n=21), promoting telemedicine and virtual visit (n=10). Five endoscopists expressed their availability to extend the working hours to perform endoscopy in the evening and during the weekend. Only 10% of the interviewed were pessimistic, fearing that the COVID-19 crisis would have not brought any changes and improvements in patient management and ED organization.

## Colorectal cancer screening

All centres claimed their availability to immediately restart the screening activity at the pre-crisis volumes, but 38 of them suggested to replace pre and post colonoscopy visits by telemedicine.

## Discussion

Present survey demonstrates that there are several barriers preventing EDs in Northern Italy to safely manage elective endoscopy out patient activity in the early recovery phase (phase-2) of pandemic. Professional societies have recently issued guidelines to safely return to elective procedures^5,6^. These guidelines call for a pre-procedure screening of patients to assess their risk of viral transmission through a structured questionnaire, preferably combining it with PCR-based testing performed within 48 hours of the procedure, the choice of PPE (in particular masks) upon the patient’s risk stratification, and policies to facilitate social distancing for patients and visitors, including appropriate spacing in the waiting and recovery rooms, restricting accompanying visitors, and distancing the procedure start times^5,6^. At present, testing for infection by RT-PCR antigen prior the procedure is not available in Italy, although this situation might change over the next months. Thus, social distancing and the appropriate us of PPE are both critical for safely resuming elective endoscopy. Unfortunately, despite 23/43 (53·5%) EDs had been recently (< 10 years) built or renovated, waiting and recovery areas are insufficiently large to guarantee the safe distance among individuals in several services, and most units are also inadequate to ensure high flows of patients, personnel and equipment in presence a such a highly transmittable infection. Other several issues may further hinder a prompt restart of elective endoscopy. Despite the current epidemic is now rapidly decelerating, endoscopy staff is still lacking in many endoscopy services, since a significant number of physicians and nurses have been either infected or are still reallocated to other departments. Moreover, some EDs rooms are still not available since converted to intensive and sub-intensive units or completely devoted to the management of infected patients. Indeed, there is a large agreement among the participants to the survey that resumption of elective endoscopy need to be accomplished very slowly, with a very limited increase in the number of procedures over the upcoming months. As a consequence, most of them considered unrealistic the possibility to return to pre-crisis workload in the next three months.

Then, this early recovery phase of pandemic is challenging: on one hand, any effort should be made to avoid an overload of EDs, on the other one a huge number of cases cancelled or postponed during the first phase of epidemic need to be rescheduled, despite the procedure lists of the upcoming months are already overbooked. In this scenario, there is an absolute need to redesign the organization models of the endoscopic units and their interaction with the territorial services. In Italy, endoscopic procedures are usually requested by GPs without a previous clinic consultation. This “open-access” endoscopy has several advantages (e.g., the reduction of costs related to endoscopy by eliminating potentially unnecessary office-based consultations, but it has also generated a very high level of inappropriateness and misuse of endoscopic resources. Up to 30% of endoscopic procedures in Italy have been judged to be inappropriate by previous surveys^7,8^. From now on, a clear and thoughtful policy regarding the timely scheduling/rescheduling of endoscopy procedures according to their clinical priority will be required. This can be achieved only by case-by-case; priority should be given to patients who have a condition that is immediately life threatening, clinically unstable, worrisome or completely intolerable and for whom even a short delay would significantly alter the patient’s prognosis. Due to the great uncertainty about the duration of the pandemic^9^, a further category of patients not to be postponed are those who do not have immediately life-threatening conditions but for whom treatment or services should not be indefinitely delayed until the end of the pandemic as for colorectal cancer screening. EDs should thus strongly consider further postponing elective, non-urgent endoscopy procedures cancellation if inappropriate (e.g. post-polypectomy surveillance colonoscopy after low risk adenoma by reviewing and categorizing the procedure lists by both prescriber physicians and endoscopists. This scenario inevitably implies, at least temporarily, the shift from an open access endoscopy to a filtered access, which promptly calls for organizational renovation of endoscopy services^10^. A prioritization model for referrals based on involvement of primary care physicians (PCPs) and specialists, which has already been successfully tested in Italy^11,12^, might represent another valuable option, albeit this process implies a close interaction between GPs and specialists, long time and resources that in this epidemic phase are difficult to find. Telemedicine, as highlighted by the present survey, could represent from now a useful tool to fill the gap between GPs and specialists, in order to improve the appropriateness of endoscopic procedures and indentify patients requiring more urgent care. Telemedicine and virtual visits have never been performed in Endoscopy Units in Italy and are presently not reimbursed by the Italian National Health System. An increase in the use of telemedicine to manage the upcoming clinic appointments is thus advisable to promote the appropriate use of resources while reducing the risk of overcrowding our services.

Measures for the return to routine endoscopy during the pandemic have been suggested by gastroenterological societies^5,6^ and experts^13–16^, but their local applicability has not been evaluated. The present survey has been conducted in Northern Italy, one of the most affected area by the COVID-19 epidemic in the world, but also the region with the most advanced heath system in Italy. It is thus conceivable that our data are generalizable to most EDs in the Western countries.

Crisis periods, like wars, are usually followed by great technological and social evolution. Nearly all endoscopists have agreed that the COVID pandemic crisis may represent a great chance to re-model and rationalize the EDs processes. Nothing will ever be the same again and what we are organizing, planning and changing today opportunity has to be exploited at best of our possibilities.

## Data Availability

I hereby confirm that all data referred to in the mauscript are available

## Contributors

GM, AR, and FR planned and designed the survey; GM and MC collected and analyzed data; GM, AR, and RF drafted the manuscript and figures. SS, CB, and FP provided critical appraisal of the manuscript. SS, CB, and FP revised the manuscript.

**Supplementary Table 1:** List of questions presented in the survey

| Characteristics of Endoscopic Units |  |
| --- | --- |
| 1 | How many procedures do you perform in your Endoscopic Unit every year?<br>A. < 5000<br>B. ≥ 5000 |
| 2 | How many physicians do you have in your Endoscopic Unit? |
| 3 | How many nurses do you have in your Endoscopic Unit? |
| 4 | How many health assistants do you have in your Endoscopic Unit? |
| 5 | When was your Endoscopic Unit built or renovated? |
| 6 | Does your Endoscopic Unit allow a differentiated clean/dirty path for the equipment? |
| 7 | How many endoscopic rooms do you have in your Unit? |
| 8 | Is your Endoscopic Unit provided with negative-pressure rooms?<br>A. Yes<br>B. No |
| Changes in your Endoscopy Unit related to the COVID-19 outbreak |  |
| 9 | **How much has the endoscopic activity of your Unit reduced?**<br>A. 100% (stopped)<br>B. 75-99%<br>C. 50-74%<br>D. 25-49%<br>E. 0-24% |
| 10 | **How many endoscopic rooms are presently not available since converted to another use?** |
| 11 | **How many physicians are presently infected?** |
| 12 | **How many physicians have been relocated to other departments?** |
| 13 | **How many nurses are presently infected?** |
| 14 | **How many nurses have been relocated to other departments?** |
| 15 | **How many health assistants are presently infected?** |
| 16 | **How many health assistants have been relocated to other departments?** |
| **Modifications in your Endoscopy Unit organization and its suitability for resuming endoscopic activity** |  |
| 17 | **In your opinion, is your Endoscopic Unit adequate to manage infected and non-infected patients?**<br>A. Yes<br>B. No<br>C. Only reducing the number of procedures |
| 18 | **Is in your Endoscopic Unit possible to have separated paths for infected and non-infected patients?**<br>A. Yes<br>B. No<br>C. Only reducing the number of procedures |
| 19 | **Is the waiting room of your Endoscopic Unit suitable to ensure adequate distance between patients/relatives/caregivers?**<br>A. Yes<br>B. No<br>C. Only reducing the number of procedures |
| 20 | **Is the recovery room of your Endoscopic Unit suitable to ensure adequate distance between patients?**<br>A. Yes<br>B. No<br>C. Only reducing the number of procedures |
| 21 | **Do you fear any shortage of PPEs in your Endoscopic Unit after resuming the endoscopic activity?**<br>A. Yes<br>B. No |
| **Perspective and proposal for resuming the endoscopic activity** |  |
| 22 | **What would you suggest to restart safely and affectively the endoscopic activity?** |
| 23 | **In your opinion, which increase in the endoscopic activity can be achieved in the month of May?**<br>A. 0% (the activity remains as today)<br>B. 10% |
|  | C. 25%<br>D. 33%<br>E. 50%<br>F. Return to the pre-COVID-19 activity |
| 24 | **In your opinion, which increase in the endoscopic activity can be achieved in the month of June?**<br>A. 0% (the activity remains as today)<br>B. 10%<br>C. 25%<br>D. 33%<br>E. 50%<br>F. Return to the pre-COVID-19 activity |
| 25 | **In your opinion, which increase in the endoscopic activity can be achieved in the month of July?**<br>A. 0% (the activity remains as today)<br>B. 10%<br>C. 25%<br>D. 33%<br>E. 50%<br>F. Return to the pre-COVID-19 activity |
| 26 | **In your opinion, when will your Endoscopic Unit return to the pre-COVID-19 activity?** |
| 27 | **Once completely re-opened, which further increase in the endoscopic activity is achievable to reduce the waiting list?**<br>A. 0% (the activity will remain as in the pre-COVID-19 period)<br>B. 10%<br>C. 25%<br>D. 33%<br>E. 50% |
| 28 | **When could the CRC screening activity restart in your Endoscopic Unit?**<br>A. Immediately (in the month of May) at the pre-COVID-19 volume<br>B. Immediately (in the month of May) at a reduced rate<br>C. I would wait to restart the screening activity |
| 29 | **In your opinion, will the COVID-19 crisis promote a significant evolution in the organization models/mentality of the Endoscopic Units?** |

